# I answer to clinical scenarios as a real doctor because I “Think”: “ChatGPTo1” new reasoning ability

**DOI:** 10.1101/2024.09.30.24314616

**Authors:** Gianluca Mondillo, Simone Colosimo, Alessandra Perrotta, Vittoria Frattolillo, Mariapia Masino, Pierluigi Marzuillo

**Affiliations:** Department of Woman, Child and of General and Specialized Surgery, Università degli Studi della Campania “Luigi Vanvitelli”, Via Luigi De Crecchio 2, Naples, Italy

**Keywords:** Artificial Intelligence, Clinical Cases, Large Language Model

## Abstract

The adoption of the ChatGPT o1 model represents a significant advancement in the management of clinical cases due to the introduction of a new structured reasoning capability, the “chain-of-thought reasoning” (CoT). In this study, 350 general medicine clinical cases were tested using ChatGPT o1 and ChatGPT o1 mini, and their performance was compared with ChatGPT 4o and ChatGPT 4o mini to evaluate diagnostic accuracy. The results showed that ChatGPT o1 achieved a correct answer rate of 93.4%, outperforming both ChatGPT 4o (82.2%) and the mini versions (ChatGPT o1 mini: 70.2% and ChatGPT 4o mini: 66.2%). The CoT technique enabled the model to provide more coherent and transparent responses, reducing the occurrence of so-called “hallucinations.” This study highlights how the ChatGPT o1 model can be a valuable tool in clinical practice, although its use requires supervision to ensure patient safety, especially in critical settings.

## Introduction

In recent years, large language models (LLMs) based on GPT, such as GPT-3 and GPT-4o, have revolutionized various sectors, including medicine, due to their ability to process large amounts of data and assist healthcare professionals in diagnosis, clinical information management, and decision-making support [1]. Despite their power, these models have shown limitations in terms of accuracy, consistency, and safety, which are crucial aspects in a sensitive field like healthcare.

With the introduction of the ‘o1’ model (OpenAI, September 12, 2024) [2], a significant step forward has been made, offering not only enhanced text processing and response generation capabilities but also a new structured reasoning methodology known as ‘chain-of-thought reasoning’ (CoT). This feature enables the model to develop a more reflective process before providing answers, thus reducing hallucinations, a phenomenon where an LLM generates inaccurate or completely erroneous responses [3]. This evolution could be particularly important in the medical field, where the accuracy of information is essential to ensuring patient well-being. Additionally, the model’s ability to resist inappropriate or harmful requests ensures greater security in managing clinical information and protecting patient privacy.

In this study, we tested 350 general medicine clinical cases using both the ChatGPT o1 model and the ChatGPT o1 mini, comparing them with results obtained from ChatGPT 4o and ChatGPT 4o mini. The goal is to assess the differences between these models in terms of diagnostic accuracy, to determine whether the technical innovations introduced by o1 represent a real advantage in clinical practice.

## Materials and Methods

For this study, we selected clinical cases from the MedQA dataset [4,5], a comprehensive collection of clinical cases commonly used for evaluating artificial intelligence models in the medical field. The selection was made randomly, by choosing the first 350 clinical cases from the dataset. Each clinical case is structured as a multiple-choice question with a single correct answer, validated by medical experts who contributed to the creation of the MedQA dataset.

The LLMs evaluated were: ChatGPT 4o, ChatGPT o1, ChatGPT o1 mini, and ChatGPT 4o mini. Each model was given the same set of clinical cases to ensure uniform testing conditions. After processing the cases, the responses provided by the models were compared with the correct answers in the dataset, already validated by human experts.

Due to the inherent limitations in the number of messages supported by the preview versions of ChatGPT o1 and ChatGPT o1 mini (30 weekly messages for o1 and 50 for o1-mini) [6], we decided to administer an average of 5 clinical cases at a time to the models. This allowed us to optimize the use of the models while adhering to the imposed limits. Consequently, we decided to analyze a total of 350 clinical cases, based on the maximum number of cases we were able to submit to the ChatGPT o1 model within its operational limitations, while still ensuring a sufficiently large sample to draw meaningful conclusions.

### Statistical analysis

For result analysis, we compared the responses obtained from the different models using contingency tables. The models’ performances were evaluated by calculating the percentage of correct answers provided by the LLMs and comparing the results with the correct answers indicated in the dataset.

P values <0.05 were considered statistically significant. Qualitative variables were compared by using chi-square test. The SciPy Python library (version 1.14.1) was used for all statistical analyses.

## Results

The results obtained from testing the 350 general medicine clinical cases showed a significant difference in the rate of correct answers between the full models (ChatGPT o1 and ChatGPT 4o) and their mini versions. ChatGPT 4o mini correctly answered 66.2% of the cases (232 out of 350), while ChatGPT o1 mini achieved a correct response rate of 70.2% (246 out of 350) (Table 1). On the other hand, the full models performed better, with ChatGPT 4o reaching 82.2% correct answers (288 out of 350) and ChatGPT o1 achieving the highest result with 93.4% correct answers (327 out of 350) (Table 1).

**Table 1.** Comparisons of correct response rate among models.

| **Models Comparisons** | **Correct Answers Model 1** | **Correct Answers Model 2** | **p-value** |
| --- | --- | --- | --- |
| **ChatGPT 4o (model 1) vs ChatGPT o1 (model 2)** | 82.2%<br>(288/350) | 93.4%<br>(327/350) | 1.68e-20 |
| **ChatGPT 4o mini (model 1) vs ChatGPT o1 mini (model 2)** | 66.2%<br>(232/350) | 70.2%<br>(246/350) | 1.13e-7 |
| **ChatGPT o1 mini (model 1) vs ChatGPT o1 (model 2)** | 70.2%<br>(246/350) | 93.4%<br>(327/350) | 4.43e-12 |
| **ChatGPT 4o mini (model 1) vs ChatGPT 4o (model 2)** | 66.2%<br>(232/350) | 82.2%<br>(288/350) | 2.47e-17 |

The confusion matrices, shown in Figure 1, display the number of correct and incorrect answers for each model.

**Figure 1.**
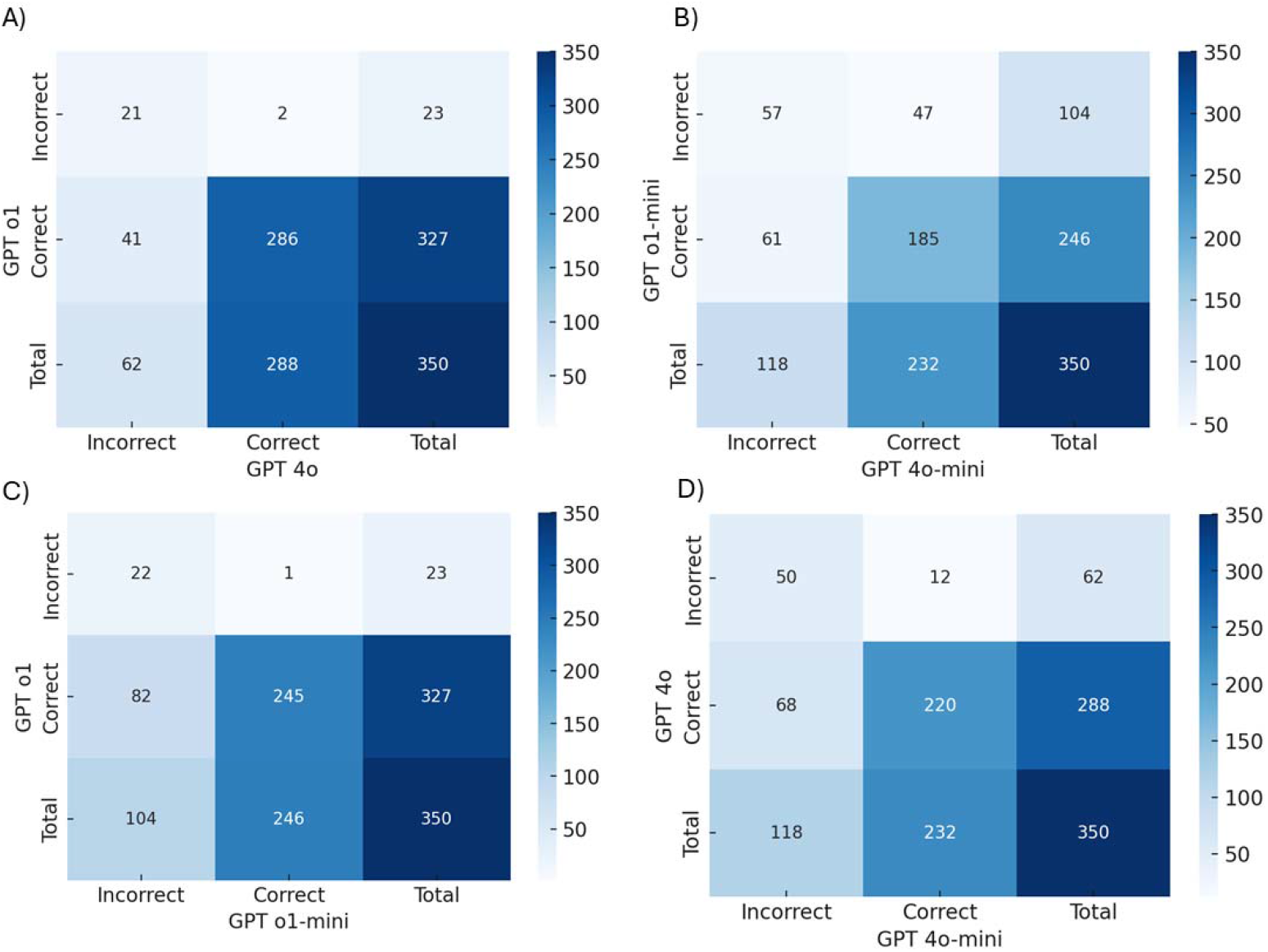
Confusion Matrices of the analyzed models. In A: ChatGPT4o vs ChatGPT o1, in B: ChatGPT o1 mini vs ChatGPT 4o mini, in C: ChatGPT o1 vs ChatGPT o1 mini, in D: ChatGPT4o vs ChatGPT 4o mini.

## Discussion

The test conducted on 350 clinical cases highlighted significant differences in performance between the models analyzed. ChatGPT o1 achieved an accuracy of 93.4%, surpassing the 82.2% obtained by ChatGPT 4o, and significantly outperforming the mini versions, with ChatGPT o1 mini at 70.2% and ChatGPT 4o mini at 66.2%. This improvement can be attributed to the use of CoT, a prompt engineering technique introduced by Wei et al. [7], which allows an LLM to break down complex problems into a series of logical steps, thereby significantly reducing the likelihood of generating incoherent or hallucinatory responses. This technique substantially contributes to improving the overall accuracy of the model’s responses.

One of the distinguishing features of ChatGPT o1 is its ability to make its reasoning process explicit, which is crucial in clinical settings where healthcare professionals need to follow and verify every logical step leading to the final answer. This transparency is particularly useful in complex diagnoses, helping to reduce the risk of diagnostic or therapeutic errors. Furthermore, the model’s ability to render its reasoning visible marks a significant step forward in Explainable AI (XAI) [8], addressing one of the main criticisms of LLMs, often perceived as ‘Black Boxes’ [9]. By providing more comprehensible and accessible explanations, these technologies can foster greater trust and integration in clinical settings. CoT not only improves the coherence of responses but also makes ChatGPT o1 particularly effective in differential diagnoses, where a sequence of clinical variables must be considered.

Although the mini versions of the LLMs analyzed showed lower performance compared to the full models, they could still play a relevant role in specific hospital settings. All OpenAI models, including the mini versions, could be integrated into clinical information systems, if properly adapted, through API (Application Programming Interface) calls [10], enabling efficient management of healthcare data and intelligent automation of repetitive tasks. The main advantage of the mini versions lies in their computational and, consequently, economic efficiency. Despite having lower accuracy, they represent an acceptable compromise for routine tasks or situations that require processing large volumes of data on a more limited budget. In these scenarios, full models, offering greater accuracy at a higher cost, should be reserved for more complex clinical tasks where precision is critical.

In the context of evaluating the safety of artificial intelligence models, OpenAI introduced the Preparedness Framework (Figure 2), a risk classification system that considers various factors, including the model’s ability to influence human behavior, its decision-making autonomy, and the risk of it being exploited in critical settings such as healthcare. This system classifies models into four risk levels: low, medium, high, and critical, depending on their potential danger [11].

**Figure 2.**
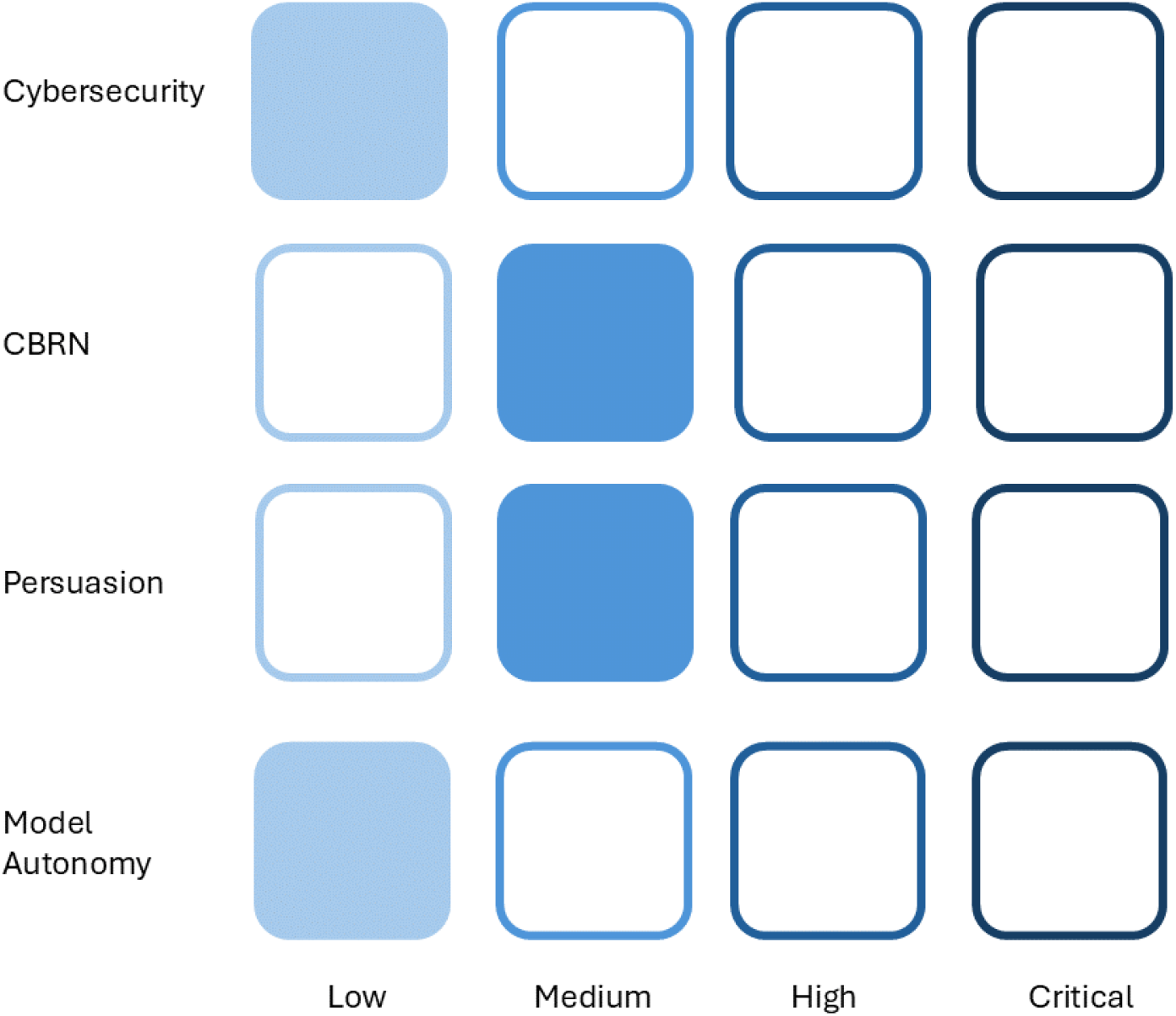
The OpenAI Preparedness Framework monitors and assesses catastrophic risks related to the development of advanced artificial intelligence models. According to the framework, the o1 models are classified as low risk for cybersecurity, as they do not exhibit significant capabilities to exploit vulnerabilities. For CBRN (chemical, biological, radiological, and nuclear) threats, the risk is considered medium, as the models can assist experts but do not enable non-experts to create threats. Persuasion is also rated as medium risk, with human-level argumentative abilities but not superior to top human writers. Finally, the risk for model autonomy is low, as the models do not exhibit capabilities for self-exfiltration or self-improvement.

ChatGPT o1 has been classified as ‘medium risk’ (ChatGPT 4o was classified as ‘low risk’) based on the combined risks from the analysis categories according to the Post-Mitigation Model Score. This evaluation reflects the fact that, despite the implementation of advanced safety measures, the model can still generate inappropriate responses under specific circumstances, such as with manipulative prompts or jailbreak attempts. Although the risks have been significantly reduced compared to previous models, this classification is particularly relevant in the clinical setting, where incorrect decisions could compromise patient safety [12]. As a result, the use of ChatGPT o1 in medical contexts requires constant supervision by healthcare professionals, especially in critical situations where the safety and reliability of responses are paramount.

Another factor to consider is that, at present, both ChatGPT o1 and ChatGPT o1 mini are not multimodal models, meaning that files cannot be uploaded through the chat interface. This limitation could reduce the model’s versatility in some clinical contexts where it would be useful to analyze documents or images for a more comprehensive evaluation.

Regarding pretraining, ChatGPT o1 was trained on a large set of public and proprietary data, including scientific and technical literature. Although the model was not specifically designed for the medical field, the variety and quality of the data it was trained on allow it to effectively manage the complexity of clinical cases. Unlike ChatGPT 4o, which was trained on multimodal data (including text, images, and audio) [13], ChatGPT o1 primarily focuses on textual data, with a specific enhancement in complex reasoning due to the integration of CoT.

In conclusion, ChatGPT o1 represents a significant advancement in the application of artificial intelligence to medicine. Its ability to provide accurate and transparent responses, supported by an explicit reasoning process, makes it a particularly valuable tool for managing complex clinical cases. A key element of this improvement is the integration of the CoT prompt, which has proven to be a powerful tool for enhancing the model’s reasoning quality by breaking down complex problems into sequential logical steps. This not only reduces the likelihood of incoherent or hallucinatory responses but also optimizes accuracy in situations where clinical decisions require more structured analysis.

Despite its advantages, the medium-risk classification of ChatGPT o1 requires a certain degree of caution in its use, especially in critical clinical contexts. Therefore, the adoption of ChatGPT o1 in the medical field demands supervised use and careful oversight to ensure patient safety and the quality of therapeutic decisions.

The mini versions of the models, while having lower performance compared to the full models, still offer a valid compromise between efficiency and cost. These versions are particularly useful in resource-constrained settings or in high-demand situations where speed of execution is crucial.

## Data Availability

All data produced in the present study are available upon reasonable request to the authors.

## Author Contributions

All the authors contributed equally.

## Institutional Review Board Statement

Not applicable, as we have not involved humans or animals.

## Informed Consent Statement

Written informed consent is not applicable.

